# Spike Protein NTD mutation G142D in SARS-CoV-2 Delta VOC lineages is associated with frequent back mutations, increased viral loads, and immune evasion

**DOI:** 10.1101/2021.09.12.21263475

**Authors:** Lishuang Shen, Timothy J. Triche, Jennifer Dien Bard, Jaclyn A. Biegel, Alexander R. Judkins, Xiaowu Gai

## Abstract

The significantly greater infectivity of the SARS-CoV-2 Delta variants of concern (VOC) is hypothesized to be driven by key mutations that result in increased transmissibility, viral load and/or evasion of host immune response. We surveyed the mutational profiles of Delta VOC genomes between September 2020 and mid-August 2021 and identified a previously unreported mutation pattern at amino acid position 142 in the N-terminal domain (NTD) of the spike protein which demonstrated multiple rounds of mutation from G142 to D142 and back. This pattern of frequent back mutations was observed at multiple time points and across Delta VOC sub-lineages. The etiology for these recurrent mutations is unclear but raises the possibility of host-directed editing of the SARS-CoV-2 genome. Within Delta VOC this mutation is associated with higher viral load, further enhanced in the presence of another NTD mutation (T95I) which was also frequently observed in these cases. Protein modeling of both mutations predicts alterations of the surface topography of the NTD by G142D, specifically disturbance of the ‘super site’ epitope that binds NTD-directed neutralizing antibodies (NAbs). The appearance of frequent and repeated G142D followed by D142G back mutations is previously unreported in SARS-CoV-2 and may represent viral adaptation to evolving host immunity characterized by increasing frequency of spike NAbs, from both prior infection and vaccine-based immunity. The emergence of alterations of the NTD in and around the main NAb epitope is a concerning development in the ongoing evolution of SARS-CoV-2 which may contribute to increased infectivity, immune evasion and ‘breakthrough infections’ characteristic of Delta VOC. Future vaccine and therapy development may benefit by recognizing the emergence of these novel spike NTD mutations and considering their impact on antibody recognition, viral neutralization, infectivity, replication, and viral load.

## Introduction

The Delta variant of concern (VOC) quickly overtook other VOCs (Alpha, Beta, Gamma, Epsilon, Lota) and by late August 2021 accounted for over 98% of new COVID-19 cases in the United States (https://covid.cdc.gov/covid-data-tracker/#variant-proportions). SARS-CoV-2 viral loads are significantly higher in both vaccinated and unvaccinated individuals infected by Delta VOC (1). At least in part this appears to be driven by increased infectivity due to the furin cleavage enhancing P681R mutation (2). Other potential mechanisms have been proposed, including reduced immune recognition resulting from diminished neutralizing antibody binding to spike protein epitopes, particularly in the receptor binding domain (RBD) of the spike protein (3).

The evolution of the SARS-CoV includes key mutations in SARS-CoV-2 proteins that interact with host immune system (4). Viral evolution and genomic diversity have also been shaped by vaccination efforts worldwide (5). Host immune response to SARS-CoV-2 infection is primarily characterized by antibodies directed against the spike protein, specifically the N-terminal domain (NTD) and the receptor-binding domain (RBD), with the latter being the focus of mRNA vaccines (6, 7). Novel mutations and new patterns of mutational evolution in the NTD and RBD domains of the spike protein may represent an evolutionary response in the context of both previous infection and worldwide vaccination efforts.

Based on analysis of available Delta VOC genomes reported from September 2020 to mid-August 2021 we identified a spike NTD mutation (G142D) in multiple major sub-lineages of Delta VOC across different ancestral mutation profiles that demonstrated an unusual pattern of frequent back mutations that frequently occurred in concert with a novel NTD mutation (T95I). The G142D mutation was associated with higher viral loads, and this effect was significantly increased by the co-occurrence of T95I.

## Materials and methods

### Ethics approval

Study design conducted at Children’s Hospital Los Angeles was approved by the Institutional Review Board under IRB CHLA-16-00429.

### SARS-CoV-2 whole genome sequencing

Whole genome sequencing of COVID-19 samples previously confirmed at Children’s Hospital Los Angeles to be positive for SARS-CoV-2 by reverse transcription-polymerase chain reaction (RT-PCR) was performed as previously described (8).

### Phylogenetic and structural analysis

Phylogenetic analysis was conducted using the NextStrain phylogenetic pipeline (version 3.0.1) (https://nextstrain.org/). Genbank NC_045512 sequence was included as SARS-CoV-2 reference genome to root the tree. Structure prediction of the hypothetical Delta spike protein with signature mutations was done by: 1) Adding the Delta spike signatures (T19R/T95I/G142D/E156G/F157del/R158del/L452R/T478K/D614G/P681R/D950N); into the fasta sequence from NC_045512.2 (reference genome) and 7C2L, S protein of SARS-CoV-2 in complex bound with antibody 4A8 (https://www.rcsb.org/structure/7C2L): 2) Predicted protein structure was derived using the Phyre2 web portal (normal mode) (9) and one-to-one threading matching using known structures: 6VSB (10), 7L2E (11), 7DK3 (12) and 7C2L (13); and 3) Alignment and superimposition of predicted structures was performed using ChimeraX Matchmaker (14): 6VSB, 7L2E, 7C2L and 7DK3.

### SARS-CoV-2 sequence and variant analysis, and emerging variant monitoring

Full-length SARS-CoV-2 sequences are annotated, curated, and monitored using a suite of bioinformatics tools, CHLA-CARD, as previously described (15-17). Statistical analysis and visualization were conducted with R (r-project/org).

## Results

### Delta VOC whole genome sequence evolutionary and phylogenetic analysis reveals recurrent G142D and T95I mutations

We analyzed SARS-CoV-2 genomes from Delta (B.1.617.2, AY.1, AY.2, AY.3-AY.12) and Kappa (B.1.617.1) VOC collected between September 2020 and August 17, 2021. Whole-genome viral sequences were obtained 1) locally by the Center for Personalized Medicine (CPM) and the Clinical Virology Laboratory, Departmental of Pathology & Laboratory Medicine at the Children’s Hospital Los Angeles, and 2) from GISAID with contributing laboratories worldwide (18, 19). All GISAID Delta VOC genomes from September 2020 to March 2021 were included. Beginning in April 2021 at least 2000 Delta VOC genomes were subsampled from the GISAID database as its expansion accelerated. The merged primary dataset consisted of 15,119 SARS-CoV-2 genomes from Delta (B.1.617.2, AY.1, AY.2, AY.3∼AY.12) and Kappa (B.1.617.1) VOCs, 12,513 of 15,119 genomes passed all the Nexstrain pipeline quality filtering and were included in the final phylogenetic analysis.

Initially and through April 2021, Delta VOCs demonstrated fewer spike protein mutations per genome than other non-Delta lineage VOCs. Beginning in May 2021, Delta VOCs established a stable level of 9.66-10.12 missense spike protein mutations per genome (**Supplemental Figure 1; Supplemental Table 1**). This generally falls with the range of spike missense mutations of non-Delta VOCs (Alpha 10.56, Beta 9.62, Gamma 12.40 in July 2021), and is greater than that observed in non-VOC lineages (7.61 in July 2021). However, Delta VOCs demonstrated a faster rate of increase in mutations per genome for genes S, ORF7a, and ORF7b, as shown in **Supplemental Figure 1**. The estimated evolutionary rate by phylogenetic analysis was 26.616 in Delta VOC, compared with 24.906 in the mixed dataset of North America, which consisted of 39.9% Delta VOC, (https://nextstrain.org/ncov/gisaid/north-america?l=clock, 2021-09-01), and 20.815 on 2021-02-25 when Delta VOC was not sampled yet.

We analyzed the distribution and evolutionary pattern of all protein-changing mutations with greater than 2% prevalence in Delta VOC. Our analysis identified Spike G142D (**Figure 1A**) and T95I (**Figure 1B**) mutations which were both recurrent and present in multiple well-separated branches or sub-lineages within the Delta VOC lineage, with prevalence rates of 55.4% and 40.1%, respectively.

**Figure 1.**
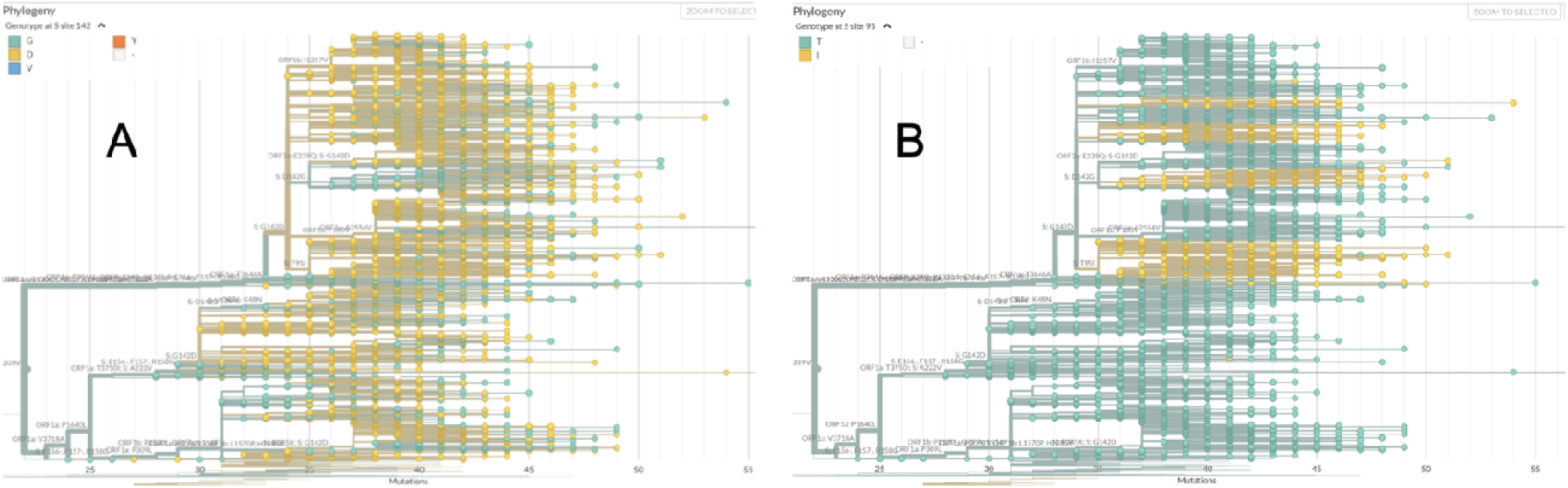
Phylogenetic analysis of the Delta and Kappa VOC genomes. **Panel A**. Delta VOC branches are colored by the amino acid at Spike protein position 142, Glycine (G) - light blue and Aspartic Acid (D) - yellow; **Panel B**. Delta VOC branches are colored by the amino acid at Spike protein position 95, Threonine (T) - light blue and Isoleucine (I) - yellow.

### G142D demonstrates frequent back mutations

Strikingly, G142D was present across almost all major Delta sub-lineages, separated from each other on the phylogenetic tree by one or more coding and non-coding mutations (**Figure 1A**). It appears that G142D was introduced into a Delta VOC major branch in late September 2020, and then again on other smaller major branches beginning in November 2020 (**Figure 1A**). Back mutation to D142G was identified in multiple branches at various time points, between early January and early July, 2021 (**Figure 1A**). This phenomenon was not observed for other spike protein mutations including T95I (**Figure 1B**). In one representative example, following the back mutation to D142G, the virus then underwent two additional protein-changing mutations (N1074S and ORD1b:P244S) and three synonymous mutations (C5239T/ORF1ab:Y1658=, A26927G/M:E135=, C13019T/ORF1ab:L4252=), and then mutated back to G142D (**Figure 2)**.

**Figure 2.**
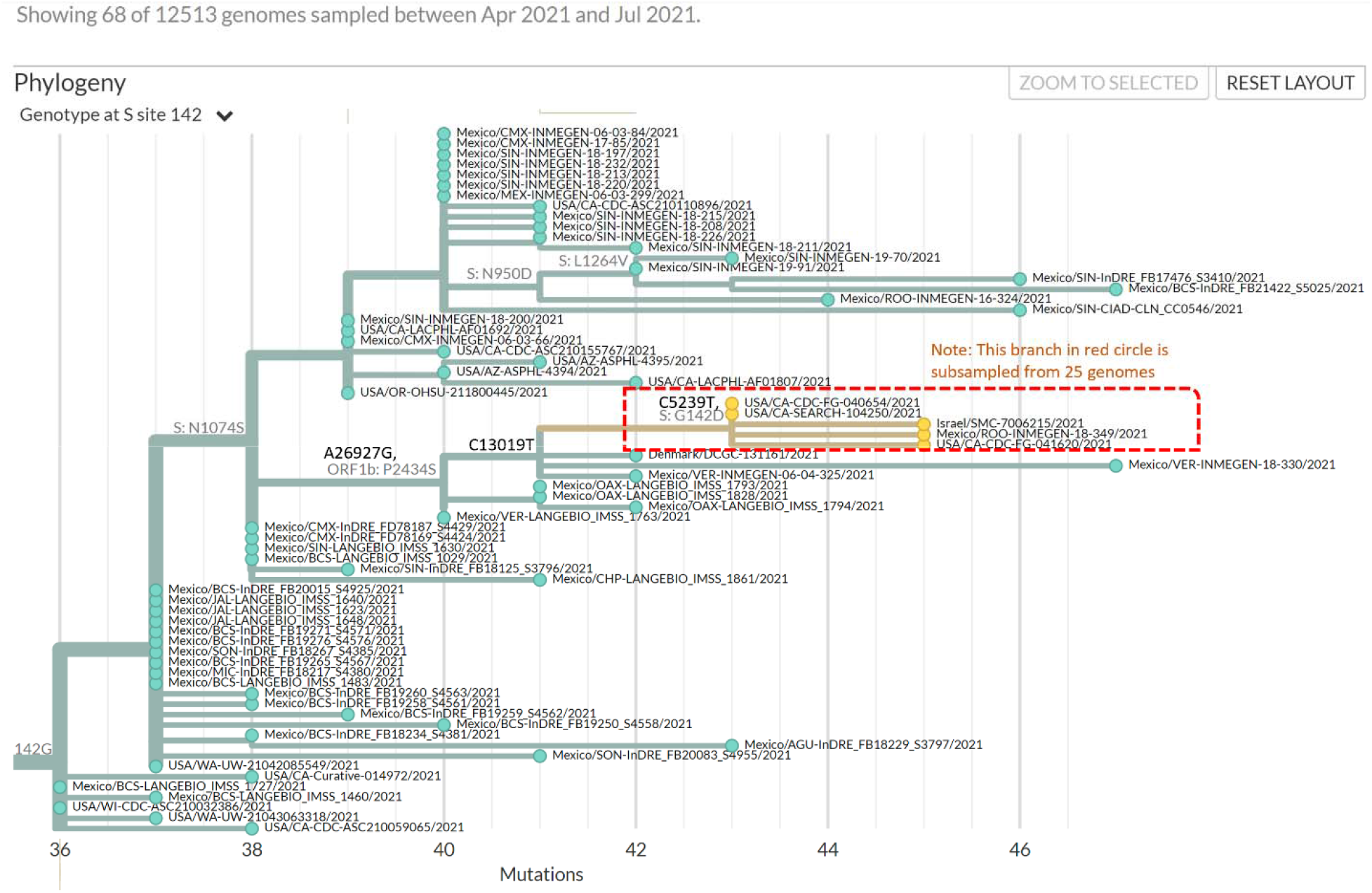
Zoomed in view of a branch of the phylogenetic tree in Figure 1.

### G142D and T95I are associated with higher viral loads within Delta VOC

Using PCR cycle threshold (Ct) values for 27,971 samples and other metadata obtained from NCBI (Bioproject Accession PRJNA731152, SARS-CoV-2 Genome sequencing and assembly – FULGENT, https://www.ncbi.nlm.nih.gov/bioproject/731152, data accessed September 8, 2021) the G142D mutation was linked to lower PCR cycle threshold (Ct) values in Delta VOC genomes. Among all Delta VOC isolates, the Ct values were significantly lower in G142D isolates than in the wild type (20.1± 3.92 vs 23.3± 3.80) (**Table 1**). The effect of G142D on the Ct values was evident across all Delta sub-lineages (**Table 2**). This effect was further enhanced by the T95I mutation which was also associated with lower Ct values across multiple sub-lineages within the Delta VOC (**Table 3**). Combined, G142D and T95I were associated with additional reduction in Ct values (18.9± 3.31 vs 20.1± 3.92) and a cumulative increase in viral load (G142D: p= 6.57e-1191 and T95I: p= 9.13e-109 by t-test). These Ct value change patterns were consistent over time (June through August) and across age and gender groups (data not shown). Using a theoretical two-fold increase per Ct cycle, these likely contribute significantly to increased Delta VOC viral loads, as much as a 9.2-fold increase for G142D alone, and up to a 21.1-fold increase when combined with T95I.

**Table 1.** Genotypes and Ct values of Delta VOC isolates collected by Fulgent Inc. (NCBI Bioproject PRJNA731152, SARS-CoV-2 Genome sequencing and assembly - FULGENT)

| **G142D** | **Spike_T95I** | **Delta VOC Lineage** | **Isolates** | **Ct value mean** |
| --- | --- | --- | --- | --- |
| Yes | Yes | Yes | 3895 | 18.9±3.31 |
| Yes | No | Yes | 22499 | 20.1±3.92 |
| No | Yes | Yes | 13** (small sample size ) | 22.0±4.04** (small sample size ) |
| No | No | Yes | 836 | 23.3±3.80 |

**Table 2:**
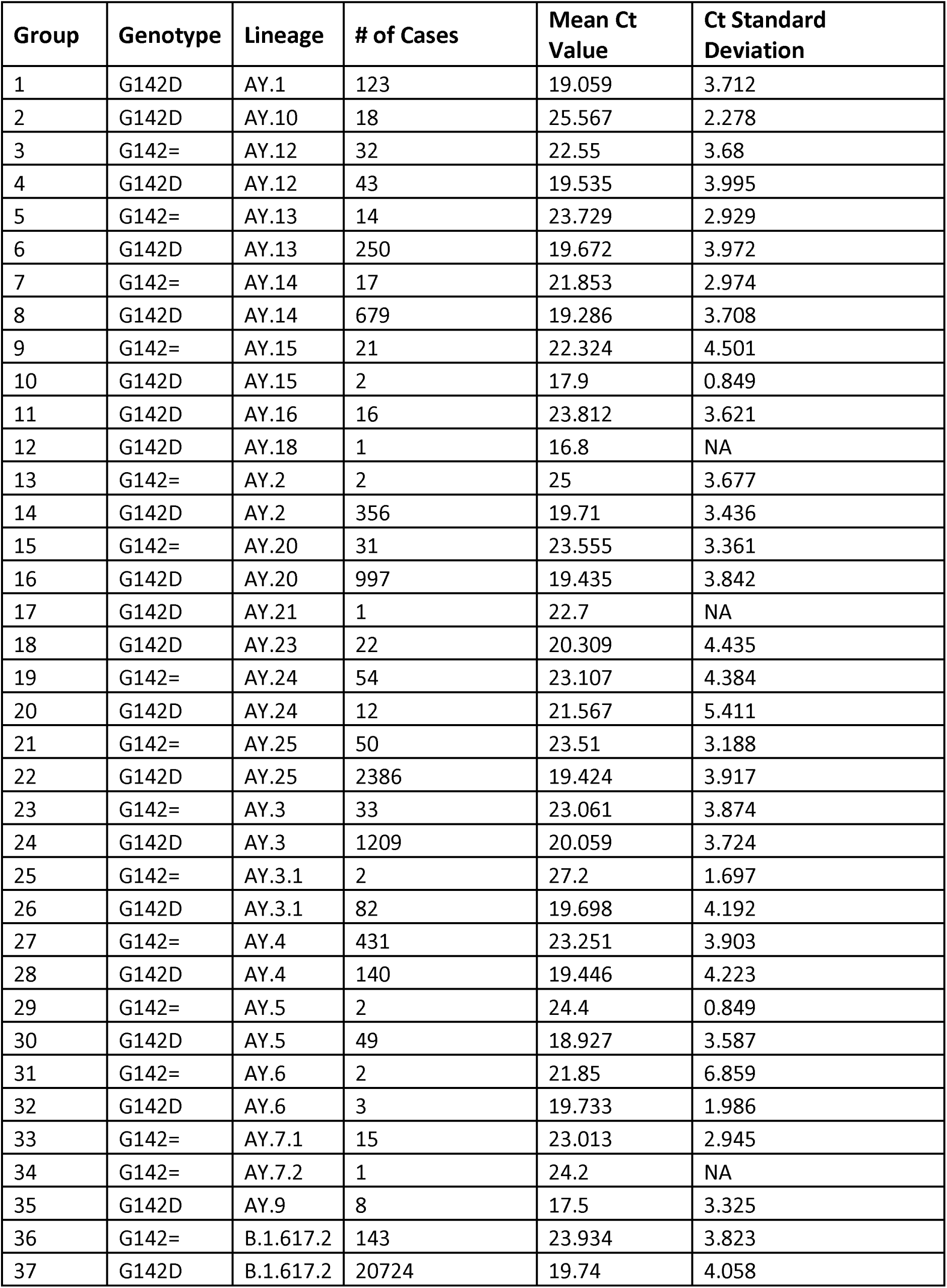
Ct values by S:G142 genotype and Pangolin lineage.

| Group | Genotype | Lineage | # of Cases | Mean Ct Value | Ct Standard Deviation |
| --- | --- | --- | --- | --- | --- |
| 1 | G142D | AY.1 | 123 | 19.059 | 3.712 |
| 2 | G142D | AY.10 | 18 | 25.567 | 2.278 |
| 3 | G142= | AY.12 | 32 | 22.55 | 3.68 |
| 4 | G142D | AY.12 | 43 | 19.535 | 3.995 |
| 5 | G142= | AY.13 | 14 | 23.729 | 2.929 |
| 6 | G142D | AY.13 | 250 | 19.672 | 3.972 |
| 7 | G142= | AY.14 | 17 | 21.853 | 2.974 |
| 8 | G142D | AY.14 | 679 | 19.286 | 3.708 |
| 9 | G142= | AY.15 | 21 | 22.324 | 4.501 |
| 10 | G142D | AY.15 | 2 | 17.9 | 0.849 |
| 11 | G142D | AY.16 | 16 | 23.812 | 3.621 |
| 12 | G142D | AY.18 | 1 | 16.8 | NA |
| 13 | G142= | AY.2 | 2 | 25 | 3.677 |
| 14 | G142D | AY.2 | 356 | 19.71 | 3.436 |
| 15 | G142= | AY.20 | 31 | 23.555 | 3.361 |
| 16 | G142D | AY.20 | 997 | 19.435 | 3.842 |
| 17 | G142D | AY.21 | 1 | 22.7 | NA |
| 18 | G142D | AY.23 | 22 | 20.309 | 4.435 |
| 19 | G142= | AY.24 | 54 | 23.107 | 4.384 |
| 20 | G142D | AY.24 | 12 | 21.567 | 5.411 |
| 21 | G142= | AY.25 | 50 | 23.51 | 3.188 |
| 22 | G142D | AY.25 | 2386 | 19.424 | 3.917 |
| 23 | G142= | AY.3 | 33 | 23.061 | 3.874 |
| 24 | G142D | AY.3 | 1209 | 20.059 | 3.724 |
| 25 | G142= | AY.3.1 | 2 | 27.2 | 1.697 |
| 26 | G142D | AY.3.1 | 82 | 19.698 | 4.192 |
| 27 | G142= | AY.4 | 431 | 23.251 | 3.903 |
| 28 | G142D | AY.4 | 140 | 19.446 | 4.223 |
| 29 | G142= | AY.5 | 2 | 24.4 | 0.849 |
| 30 | G142D | AY.5 | 49 | 18.927 | 3.587 |
| 31 | G142= | AY.6 | 2 | 21.85 | 6.859 |
| 32 | G142D | AY.6 | 3 | 19.733 | 1.986 |
| 33 | G142= | AY.7.1 | 15 | 23.013 | 2.945 |
| 34 | G142= | AY.7.2 | 1 | 24.2 | NA |
| 35 | G142D | AY.9 | 8 | 17.5 | 3.325 |
| 36 | G142= | B.1.617.2 | 143 | 23.934 | 3.823 |
| 37 | G142D | B.1.617.2 | 20724 | 19.74 | 4.058 |

**Table 3:** Ct values by S:T95 genotype and Pangolin lineage.

| **Group** | **Genotype** | **Lineage** | **# of Cases** | **Mean Ct Value** | **Ct Standard Deviation** |
| --- | --- | --- | --- | --- | --- |
| 1 | T95= | AY.1 | 90 | 20.076 | 3.638 |
| 2 | T95I | AY.1 | 33 | 16.288 | 2.213 |
| 3 | T95= | AY.10 | 18 | 25.567 | 2.278 |
| 4 | T95= | AY.12 | 65 | 21.006 | 4.036 |
| 5 | T95I | AY.12 | 10 | 19.62 | 4.686 |
| 6 | T95= | AY.13 | 264 | 19.887 | 4.023 |
| 7 | T95= | AY.14 | 696 | 19.349 | 3.711 |
| 8 | T95= | AY.15 | 21 | 22.238 | 4.508 |
| 9 | T95I | AY.15 | 2 | 18.8 | 3.677 |
| 10 | T95= | AY.16 | 16 | 23.812 | 3.621 |
| 11 | T95= | AY.18 | 1 | 16.8 | NA |
| 12 | T95= | AY.2 | 358 | 19.739 | 3.454 |
| 13 | T95= | AY.20 | 187 | 23.325 | 3.798 |
| 14 | T95I | AY.20 | 841 | 18.722 | 3.385 |
| 15 | T95= | AY.21 | 1 | 22.7 | NA |
| 16 | T95= | AY.23 | 21 | 20.019 | 4.326 |
| 17 | T95I | AY.23 | 1 | 26.4 | NA |
| 18 | T95= | AY.24 | 66 | 22.827 | 4.581 |
| 19 | T95= | AY.25 | 2436 | 19.508 | 3.945 |
| 20 | T95= | AY.3 | 1240 | 20.137 | 3.758 |
| 21 | T95I | AY.3 | 2 | 21.2 | 4.384 |
| 22 | T95= | AY.3.1 | 84 | 19.876 | 4.302 |
| 23 | T95= | AY.4 | 460 | 23.172 | 3.955 |
| 24 | T95I | AY.4 | 111 | 18.782 | 3.879 |
| 25 | T95= | AY.5 | 51 | 19.141 | 3.677 |
| 26 | T95= | AY.6 | 5 | 20.58 | 3.883 |
| 27 | T95= | AY.7.1 | 14 | 23.257 | 2.895 |
| 28 | T95I | AY.7.1 | 1 | 19.6 | NA |
| 29 | T95= | AY.7.2 | 1 | 24.2 | NA |
| 30 | T95= | AY.9 | 8 | 17.5 | 3.325 |
| 31 | T95= | B.1.617.2 | 17811 | 19.966 | 4.129 |
| 32 | T95I | B.1.617.2 | 3056 | 18.623 | 3.507 |

### G142D and T95I likely alter NTD surface topography in the antibody binding region

Phyre predicted secondary structure shows that G142D plus T95I are associated with the potential conversion of a β-strand to α-helix structure around aa159-167 and aa183-190. The Delta’s major NTD region 3-D structural difference is around aa150-158, close to the G142D and E156G+157_158del. This change in surface topography extends up and into the N3-N5 loop antibody binding ‘super site’. The combination G142D and T95I with other Delta VOC alterations, including aa157_158del and substitutions of E156G, is predicted to alter the original 4-8 (7L2E) and 4A48 (7C2L) antibody binding epitope, diminishing antibody binding to this region. Among them, new antibody facing residues are likely exposed by this change in conformation, including TYR145, LYS150, SER151, TRP152 ASP176, GLU178, which are predicted to diminish binding by repelling antibody binding. Thus, the G142D and T95I mutations are predicted to alter the surface topography of the NTD, including the antibody binding “super site” formed by the N3 and N5 loop structures (**Figure 3)**. Within Delta, the G142D further caused TRP152 position to shift outwards to the antibody 41-8 side by 10.786 Å than Delta without G142D, and by 13.457Å than non-Delta template 7L2E (**Figure 3**). Missense3D predicted a 186 Å^3 enlargement of the pocket at T95I, yet T95I is not changing the topology as significantly as G142D which is not surprising for a buried residue. In the absence of both mutations, the β-strand is conserved (**Supplemental Figure 2**). Interestingly, if only T95I is absent, the native β-strand is predicted to be converted to a disorganized coil around aa159-165, but the aa183-190 region remains α-helical. All predictions based on these NTD mutations had the prominent protrusion of a loop structure in the furin cleavage site around aa681-685, associated with the P681R mutation, thus retaining the enhanced protease cleavage of S1 from S2 associated with this mutation (data not shown).

**Figure 3.**
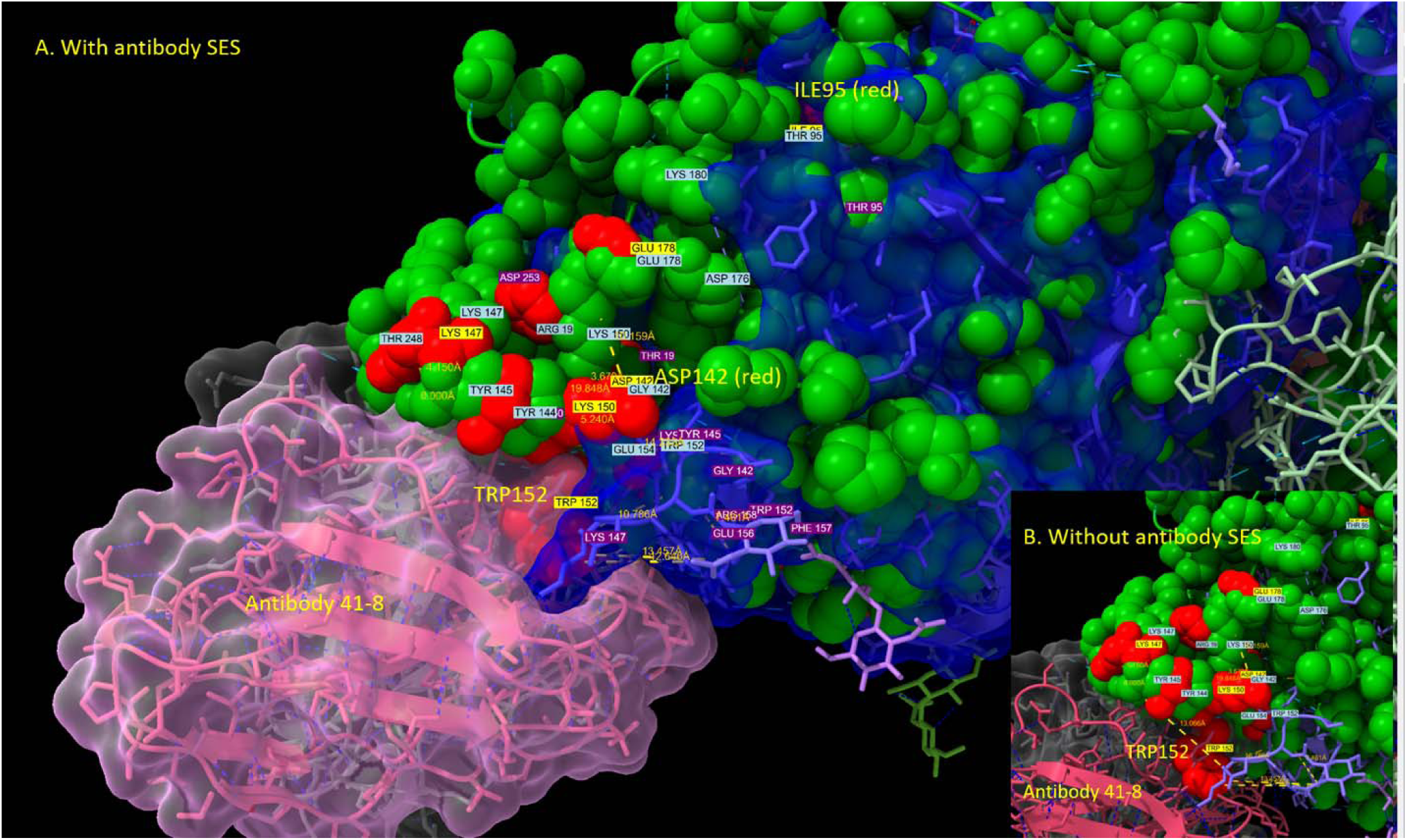
Superimposed 7L2E and Delta mutant spike protein structure at the NTD region predicts altered surface topography unfavorable to NAb binding. *De novo* predicted NTD structure of Delta mutant with both G142D and T95I: residues in red, residue labels in yellow boxes; Delta mutant without G142D or T95I: residues in green and residue labels in light blue boxes. 7L2E: spike residues in orange, residue labels in purple boxes and the solvent excluded surface (SES) in blue. TRP152 residue in the *De novo* predicted structure of the Delta variant with both G142D and T95I clashes with 7L2E antibody, which is colored in red with and without (inset) the solvent excluded surface (SES) shown in light pink bubble).

## Discussion

Through comprehensive phylogenetic analysis, mutation profiling, and public database mining, we identified a recurrent and frequently repeated back mutation at spike protein G142 in the NTD region of the spike protein. The repeated gain, loss, and regain of this mutation at the same position across multiple sub-lineages has not to our knowledge been reported in SARS-CoV-2 or other coronaviruses. The basis for this change is unclear but the mutational pattern is inconsistent with the previously observed fidelity rate of SARS-CoV-2 polymerase, which is enhanced by nsp14 ExoN proofreading (20). However, it seems likely that random RNA polymerase errors changes are not the only driver of changes in the SARS-CoV-2 genome. Mutational surveys of published SARS-CoV-2 genomes reveal overrepresentations of nucleotide changes bearing the signature of RNA editing: adenosine-to-inosine (A-G) changes from ADAR deaminases and cytosine-to-uracil (C-U) changes from APOBEC deaminases (21-24). The role of APOBEC and ADAR proteins in host innate immunity and their viral directed activities raises the possibility of host immunity contributing to the intra-host heterogeneity observed in the SARS-CoV-2 genome (22, 24-26). Indeed, this has been proposed as the underlying mechanism for the D614G mutation (27). Whether the observed frequent mutation and back mutation events at position 142 reported here are due to RNA editing is of interest and requires analysis beyond the scope of this study.

Delta VOC cases with spike NTD mutation G142D demonstrated increased viral loads, an effect that was further enhanced in the presence of the second NTD mutation (T95I) this study. Precisely how these changes lead to higher viral loads and perhaps contribute to increased transmission in the Delta VOC remains to be fully elucidated. However, our results suggest that reduced NAb binding to the NTD is one potential mechanism that deserves further investigation. The G142D mutation is located in close proximity to the alterations of E156G and DEL157-158 reported in all Delta VOC that is part of the N3 loop of the NTD ‘super site’ epitope recognized by NTD-directed NAbs and associated with reduced viral neutralization (28-32). Despite the well-known challenges of *in-silico* protein structure e prediction on the spike protein (e.g., pre- and post-fusion conformation, the impact of D614G and P681R mutations on furin cleavage structure, and native spike conformation vs various mutated spike protein and their effect on antibody binding to different NTD or RBD epitopes) all of our structural predictions modelled against different templates (native, NTD- or RBD antibody-bound) consistently predicted that G142D and T95I mutations contribute to significant changes in the NTD secondary structure, including the potential conversion of β-strand to α-helix around aa159-167 and 183-190 regions, hydrogen-bond changes due to altered residue charge at G142D, an enlarged pocket at T95I, and overall three dimensional NTD surface topography that is unfavorable to NAb binding in the presence of other Delta VOC signature mutations. It is of interest that the most recently identified VOC (Mu variant, B.1.621) differs from previous VOCs primarily by additional mutations immediately adjacent to the G142D reported here (i.e., Y144T, Y145S, 146N), further implicating this region of the NTD as important to immune escape from NTD directed Nabs. (33)

The increased frequency of Spike mutations, especially in the NTD, in the recently emergent Delta and Mu VOCs has not been observed in previous VOCs. Additional sequence data collection and analysis will be required to determine if this is an example of host-cell induced RNA editing or other mechanism that results in rapid generation of multiple variants, from which those that are biologically more ‘fit’ can emerge and re-emerge (21-23). Whatever the underlying mechanism, this change raises important questions about the evolutionary trajectory of SARS-CoV-2 with potentially significant clinical implications for reinfections and vaccine breakthrough infections which have become hallmarks of the Delta VOC. The combination of diminished immune recognition in both the RBD and NTD, here linked to a recurring mutation at G142D, in concert with a T95I mutation, raises the question of whether the observed mutations of the Delta VOC may at least in part reflect evolutionary adaptation to the growing percentage of the population immune to prior variants, whether by previous infection or vaccinations that primarily target the spike RBD, as this pandemic continues. If so, increased vigilance will be required to detect emergence of new NTD region mutations that may confer even greater immune evasion, replication, and transmission advantages to future Delta sub-lineages and SARS-CoV-2 VOCs yet to emerge.

## Data Availability

SARS-CoV-2 genome sequences used in the study were obtained from GISAID directly or from positive COVID-19 samples sequenced by the Children's Hospital Los Angeles (CHLA). CHLA SARS-CoV-2 sequences have also been submitted to GISAID.

## Acknowledgments

We would like to acknowledge all members of the Department of Pathology and Laboratory Medicine (PLM) at Children’s Hospital Los Angeles for dedication towards providing excellent patient care throughout the pandemic, including especially the rapid launch of both the COVID-19 diagnostic test in the Clinical Microbiology and Virology Laboratory and the SARS-CoV-2 whole genome sequencing assay at the Center for Personalized Medicine in March 2020, both with the support of Thermo Fisher and Paragon Genomics, with whom these assays were developed. We would like to acknowledge the frontline healthcare workers who remain devoted in the fight against COVID-19. We would also like to acknowledge all institutions that have contributed the SARS-CoV-2 sequences timely, as well as NCBI, GISAID, and Nextstrain for providing valuable resources for SARS-CoV-2 genomics.

## Online material

Video: http://34.211.4.186/iva/tmp/Delta_core_mutation_with_T95I_G614D_labelled.mp4

---

**Supplemental Figure 1.**
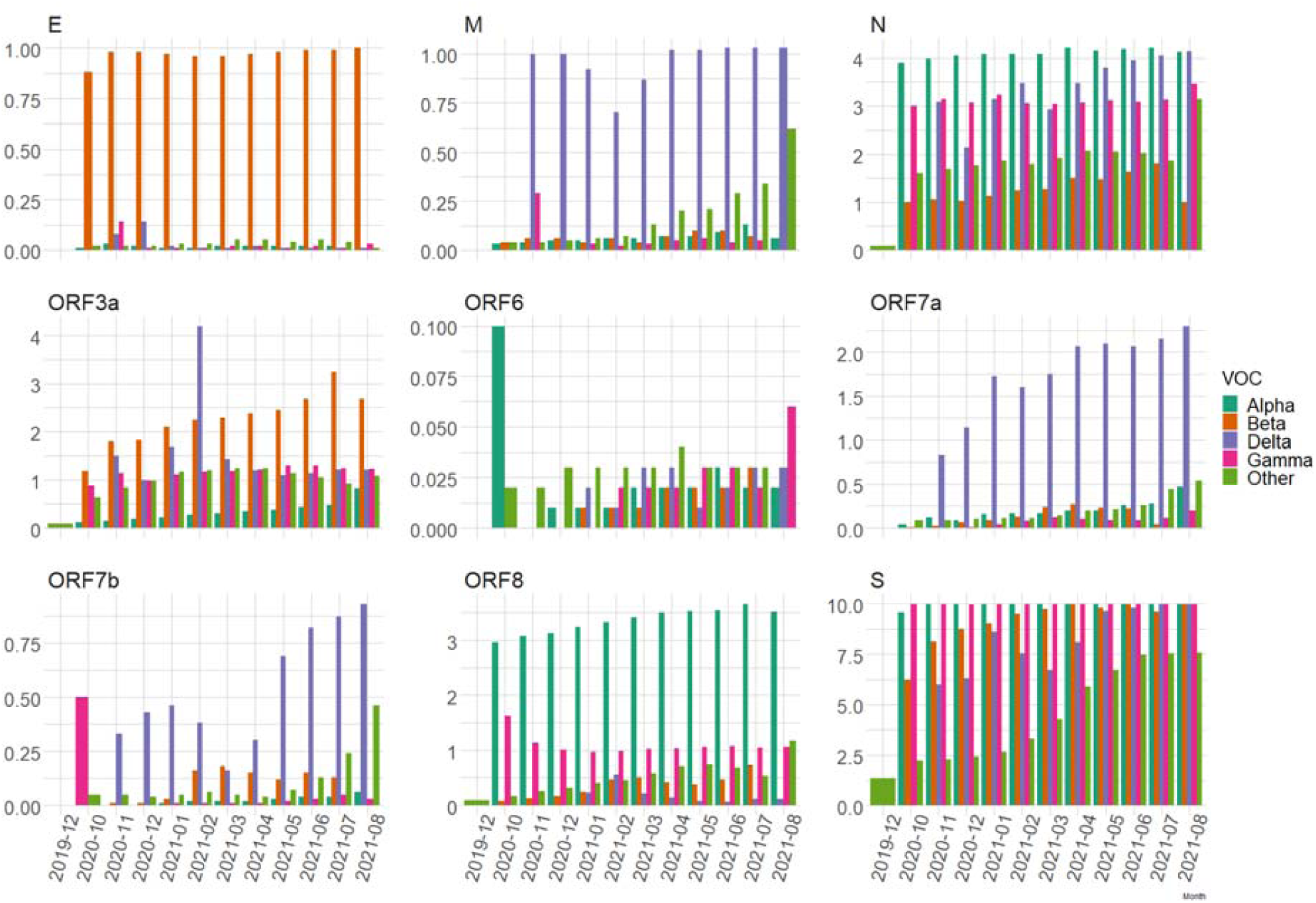
Mutations per gene per genome in different lineages.

**Supplemental Figure 2.**
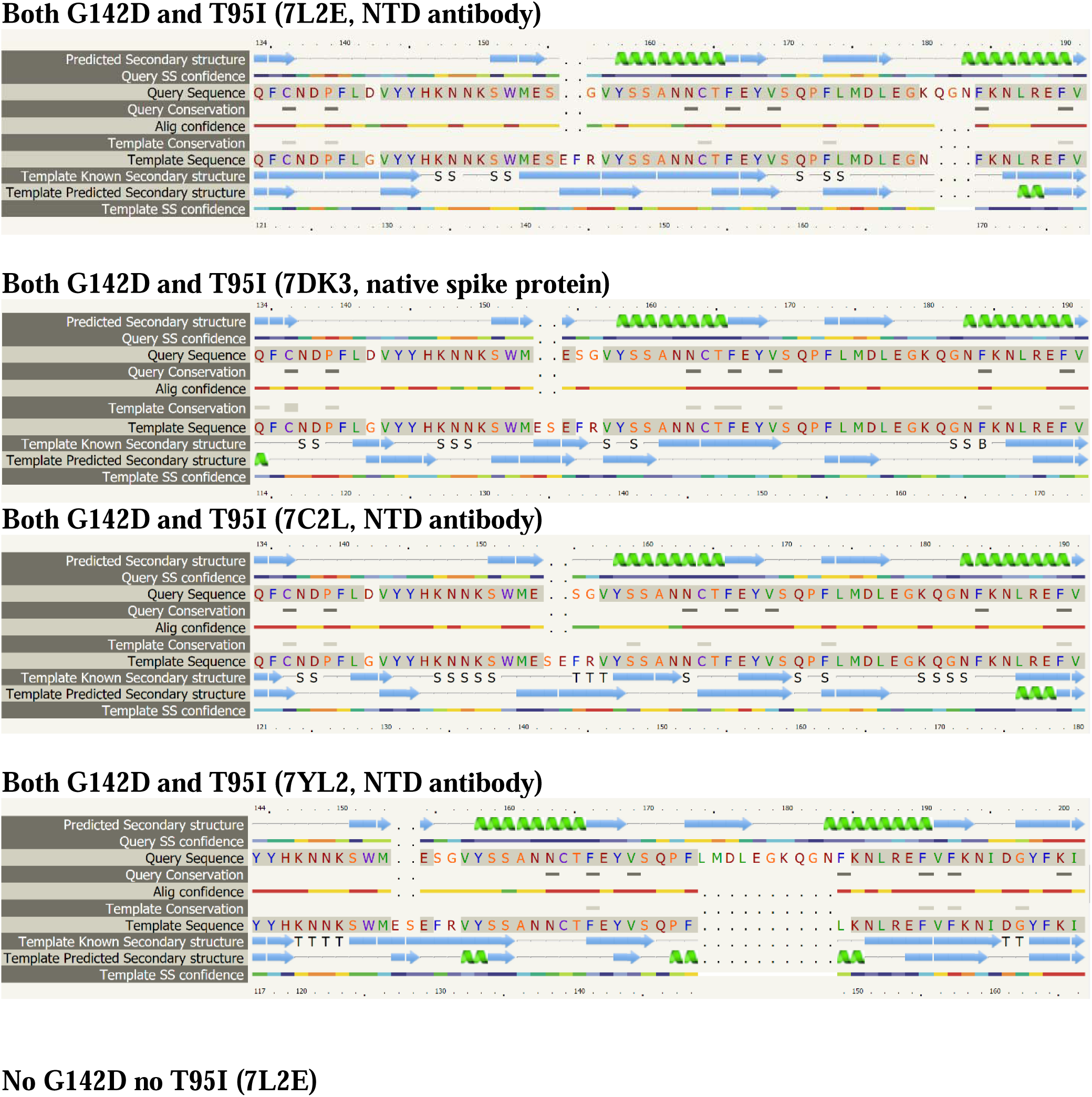
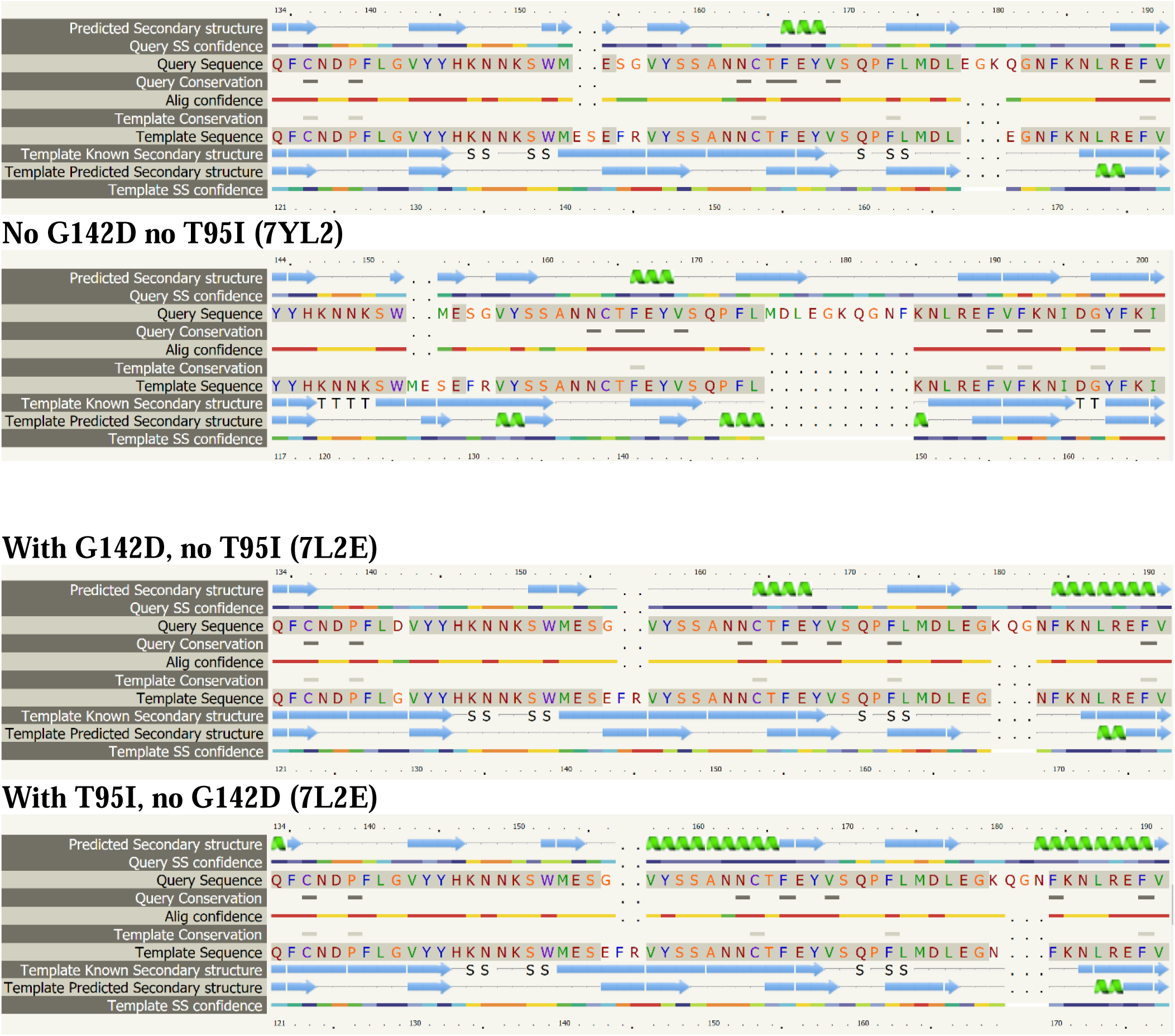
Phyre predicted secondary structures of Delta core mutations through one to one threading onto 7L2E, 7C2L, 7YL2 as templates. The prediction is 3-state: either α-helix, β-strand or coil. Green helices represent α-helices, blue arrows indicate β-strands and faint lines indicate coil. The ‘SS confidence’ line indicates the confidence in the prediction, with red being high confidence and blue low confidence.

**Supplemental Table 1.**
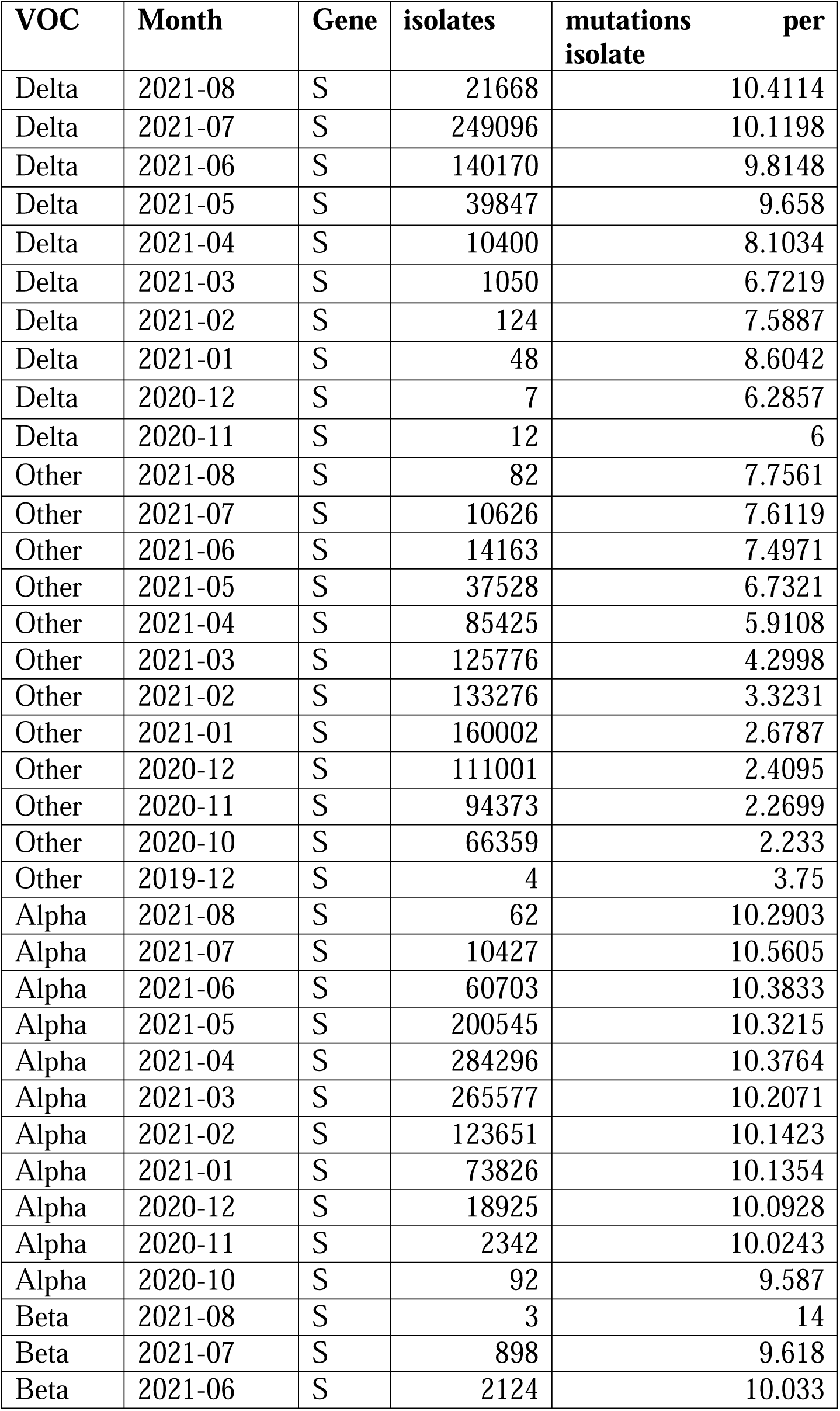
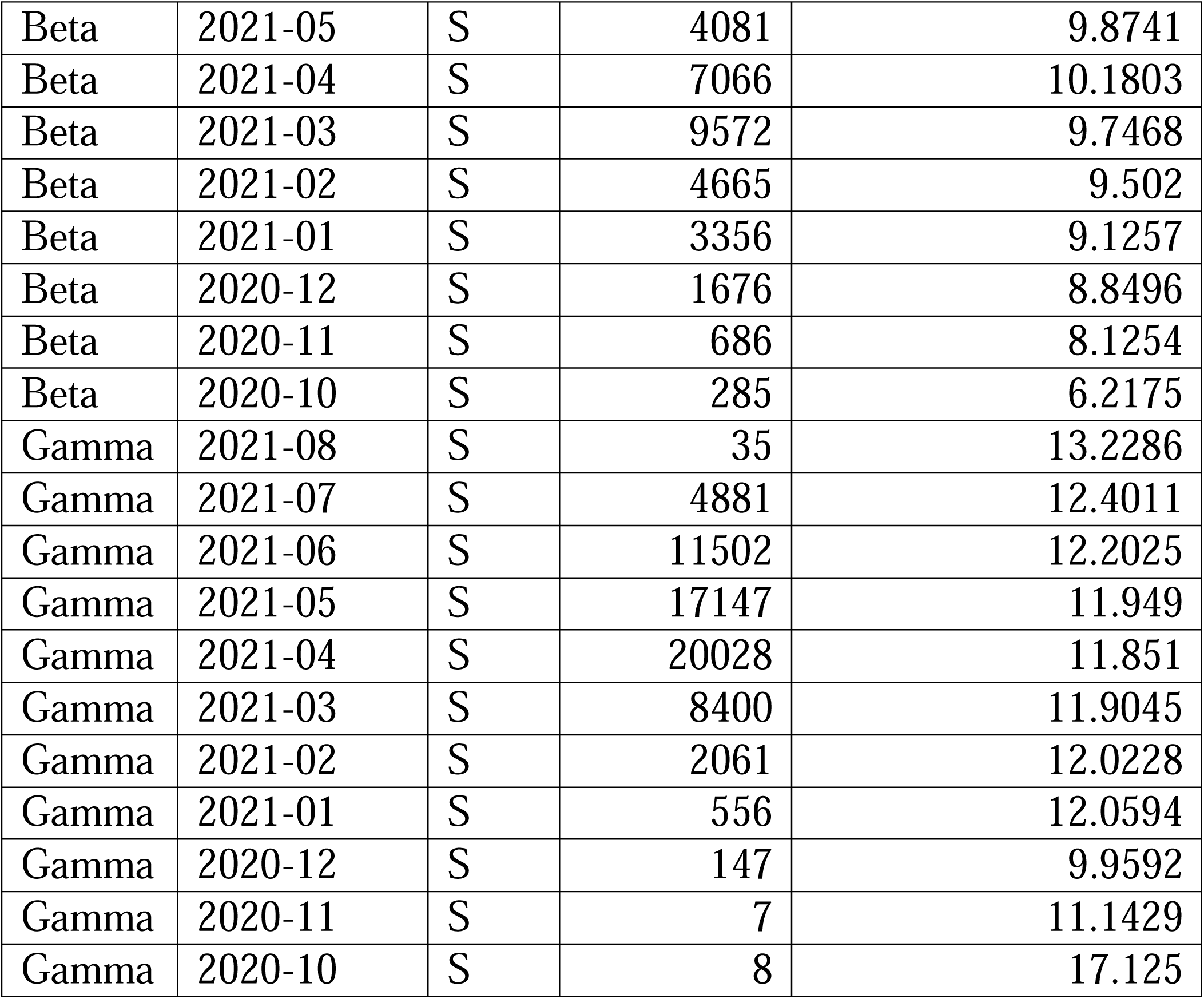
Number of Spike mutations per isolate in different VOC lineages and at different times.

## References

1. Morris P, Sachithanandham J, Amadi A, Gaston D, Li M, Swanson NJ, Schwartz M, Klein EY, Pekosz A, Mostafa HH. Infection with the SARS-CoV-2 Delta Variant is Associated with Higher Infectious Virus Loads Compared to the Alpha Variant in both Unvaccinated and Vaccinated Individuals. medRxiv 2021.08.15.21262077; doi: https://doi.org/10.1101/2021.08.15.21262077

2. Liu Y, Liu J, Johnson BA, Xia H, Ku Z, Schindewolf C, Widen SG, An Z, Weaver SC, Menachery VD, Xie X, Shi PY. Delta spike P681R mutation enhances SARS-CoV-2 fitness over Alpha variant. bioRxiv 2021.08.12.456173; doi: https://doi.org/10.1101/2021.08.12.456173.

3. Planas D, Veyer D, Baidaliuk A, Staropoli I, Guivel-Benhassine F, Rajah MM, Planchais C, Porrot F, Robillard N, Puech J, Prot M, Gallais F, Gantner P, Velay A, Le Guen J, Kassis-Chikhani N, Edriss D, Belec L, Seve A, Courtellemont L, Péré H, Hocqueloux L, Fafi-Kremer S, Prazuck T, Mouquet H, Bruel T, Simon-Lorière E, Rey FA, Schwartz O. Reduced sensitivity of SARS-CoV-2 variant Delta to antibody neutralization. Nature. 2021 Aug;596(7871):276–280. doi: 10.1038/s41586-021-03777-9. Epub 2021 Jul 8. PMID: 34237773.

4. Banoun H. Evolution of SARS-CoV-2: Review of Mutations, Role of the Host Immune System. Nephron. 2021;145(4):392–403. doi: 10.1159/000515417. Epub 2021 Apr 28. PMID: 33910211; PMCID: PMC8247830.

5. Niesen MJM, Anand P, Silvert E, Suratekar R, Pawlowski C, Chosh P, Lenehan P, Hughes T, Zemmour D, O’Horo JC, Yao JD, Pritt BS, Norgan A, Hurt RT, Badley AD, Venkatakrishnan AJ, Soundararajan V. COVID-19 vaccines dampen genomic diversity of SARS-CoV-2: Unvaccinated patients exhibit more antigenic mutational variance. medRxiv 2021.07.01.21259833; doi: 10.1101/2021.07.01.21259833

6. Williams TC, Burgers WA. SARS-CoV-2 evolution and vaccines: cause for concern? Lancet Respir Med. 2021 Apr;9(4):333–335. doi: 10.1016/S2213-2600(21)00075-8. Epub 2021 Jan 29. PMID: 33524316; PMCID: PMC8009632.

7. Greaney AJ, Loes AN, Crawford KHD, Starr TN, Malone KD, Chu HY, Bloom JD. Comprehensive mapping of mutations in the SARS-CoV-2 receptor-binding domain that affect recognition by polyclonal human plasma antibodies. Cell Host Microbe. 2021 Mar 10;29(3):463-476.e6. doi: 10.1016/j.chom.2021.02.003. Epub 2021 Feb 8. PMID: 33592168; PMCID: PMC7869748.

8. Pandey U, Yee R, Shen L, Judkins AR, Bootwalla M, Ryutov A. Maglinte DT, Ostrow D, Precit M, Biegel JA, Bender JM, Gai X, Dien Bard J. High prevalence of SARS-CoV-2 genetic variation and D614G mutation in pediatric patients with COVID-19. Open Forum Infectious Diseases, ofaa551.

9. Kelley LA, Mezulis S, Yates CM, Wass MN, Sternberg MJ. The Phyre2 web portal for protein modeling, prediction and analysis. Nat Protoc. 2015 Jun;10(6):845–58. doi: 10.1038/nprot.2015.053. Epub 2015 May 7. PMID: 25950237; PMCID: PMC5298202.

10. Wrapp D, Wang N, Corbett KS, Goldsmith JA, Hsieh CL, Abiona O, Graham BS, McLellan JS. Cryo-EM structure of the 2019-nCoV spike in the prefusion conformation. Science. 2020 Mar 13;367(6483):1260–1263. doi: 10.1126/science.abb2507. Epub 2020 Feb 19. PMID: 32075877; PMCID: PMC7164637.

11. Cerutti G, Guo Y, Zhou T, Gorman J, Lee M, Rapp M, Reddem ER, Yu J, Bahna F, Bimela J, Huang Y, Katsamba PS, Liu L, Nair MS, Rawi R, Olia AS, Wang P, Zhang B, Chuang GY, Ho DD, Sheng Z, Kwong PD, Shapiro L. Potent SARS-CoV-2 neutralizing antibodies directed against spike N-terminal domain target a single supersite. Cell Host Microbe. 2021 May 12;29(5):819-833.e7. doi: 10.1016/j.chom.2021.03.005. Epub 2021 Mar 12. PMID: 33789084; PMCID: PMC7953435.

12. Xu C, Wang Y, Liu C, Zhang C, Han W, Hong X, Wang Y, Hong Q, Wang S, Zhao Q, Wang Y, Yang Y, Chen K, Zheng W, Kong L, Wang F, Zuo Q, Huang Z, Cong Y. Conformational dynamics of SARS-CoV-2 trimeric spike glycoprotein in complex with receptor ACE2 revealed by cryo-EM. Sci Adv. 2021 Jan 1;7(1):eabe5575. doi: 10.1126/sciadv.abe5575. PMID: 33277323; PMCID: PMC7775788.

13. Chi X, Yan R, Zhang J, Zhang G, Zhang Y, Hao M, Zhang Z, Fan P, Dong Y, Yang Y, Chen Z, Guo Y, Zhang J, Li Y, Song X, Chen Y, Xia L, Fu L, Hou L, Xu J, Yu C, Li J, Zhou Q, Chen W. A neutralizing human antibody binds to the N-terminal domain of the Spike protein of SARS-CoV-2. Science. 2020 Aug 7;369(6504):650–655. doi: 10.1126/science.abc6952. Epub 2020 Jun 22. PMID: 32571838; PMCID: PMC7319273.

14. Pettersen EF, Goddard TD, Huang CC, Meng EC, Couch GS, Croll TI, Morris JH, Ferrin TE. UCSF ChimeraX: Structure visualization for researchers, educators, and developers. Protein Sci. 2021 Jan;30(1):70–82. doi: 10.1002/pro.3943. Epub 2020 Oct 22. PMID: 32881101; PMCID: PMC7737788.

15. Shen L, Maglinte DT, Ostrow D, Pandey U, Bootwalla M, Ryutov A, Govindarajan A, Ruble D, Han J, Triche TJ, Dien Bard J, Biegel JA, Judkins AR, Gai X. Children’s Hospital Los Angeles COVID-19 Analysis Research Database (CARD) - A Resource for Rapid SARS-CoV-2 Genome Identification Using Interactive Online Phylogenetic Tools. bioRxiv 2020.05.11.089763; doi: 10.1101/2020.05.11.089763

16. Shen L, Bard JD, Triche TJ, Judkins AR, Biegel JA, Gai X. Emerging variants of concern in SARS-CoV-2 membrane protein: a highly conserved target with potential pathological and therapeutic implications. Emerg Microbes Infect. 2021 Dec;10(1):885–893. doi: 10.1080/22221751.2021.1922097. PMID: 33896413; PMCID: PMC8118436.

17. Shen L, Bard JD, Triche TJ, Judkins AR, Biegel JA, Gai X. Rapidly emerging SARS-CoV-2 B.1.1.7 sub-lineage in the United States of America with spike protein D178H and membrane protein V70L mutations. Emerg Microbes Infect. 2021 Dec;10(1):1293–1299. doi: 10.1080/22221751.2021.1943540. PMID: 34125658; PMCID: PMC8238060.

18. Elbe S, Buckland-Merrett G. Data, disease and diplomacy: GISAID’s innovative contribution to global health. Glob Chall. 2017 Jan 10;1(1):33–46. doi:10.1002/gch2.1018. PMID: 31565258; PMCID: PMC6607375

19. Shu Y, McCauley J. GISAID: Global initiative on sharing all influenza data - from vision to reality. Euro Surveill. 2017 Mar 30;22(13):30494. doi: 10.2807/1560-7917.ES.2017.22.13.30494. PMID: 28382917; PMCID: PMC5388101.

20. Yan L, Yang Y, Li M, Zhang Y, Zheng L, Ge J, Huang YC, Liu Z, Wang T, Gao S, Zhang R, Huang YY, Guddat LW, Gao Y, Rao Z, Lou Z. Coupling of N7-methyltransferase and 3’-5’ exoribonuclease with SARS-CoV-2 polymerase reveals mechanisms for capping and proofreading. Cell. 2021 Jun 24;184(13):3474-3485.e11. doi: 10.1016/j.cell.2021.05.033. Epub 2021 May 24. PMID: 34143953; PMCID: PMC8142856.

21. Di Giorgio S, Martignano F, Torcia MG, Mattiuz G, Conticello SG. Evidence for host-dependent RNA editing in the transcriptome of SARS-CoV-2. Sci Adv. 2020 Jun 17;6(25):eabb5813. doi: 10.1126/sciadv.abb5813. PMID: 32596474; PMCID: PMC7299625.

22. Wang R, Hozumi Y, Zheng YH, Yin C, Wei GW. Host Immune Response Driving SARS-CoV-2 Evolution. Viruses. 2020 Sep 27;12(10):1095. doi: 10.3390/v12101095. PMID: 32992592; PMCID: PMC7599751.

23. Mourier T, Sadykov M, Carr MJ, Gonzalez G, Hall WW, Pain A. Host-directed editing of the SARS-CoV-2 genome. Biochem Biophys Res Commun. 2021 Jan 29;538:35–39. doi: 10.1016/j.bbrc.2020.10.092. Epub 2020 Nov 5. PMID: 33234239; PMCID: PMC7643664.

24. Pathak AK, Fatihi S, Abbas T, Uppili B, Mishra GP, Ghosh A, Banu S, Bhoyar RC, Jain A, Divakar MK, Imran M, Faruq M, Sowpati DT, Raghav SK, Thukral L, Mukerji M. Potential role of host-mediated RNA editing in intra-host variability of SARS-CoV-2 genomes. bioRxiv 2020.12.09.417519; doi: https://doi.org/10.1101/2020.12.09.417519

25. Samuel CE. ADARs: viruses and innate immunity. Curr Top Microbiol Immunol. 2012;353:163–95. doi: 10.1007/82_2011_148. PMID: 21809195; PMCID: PMC3867276.

26. Samuel CE. Adenosine deaminases acting on RNA (ADARs) are both antiviral and proviral. Virology. 2011 Mar 15;411(2):180–93. doi: 10.1016/j.virol.2010.12.004. Epub 2011 Jan 5. PMID: 21211811; PMCID: PMC3057271.

27. Vlachogiannis NI, Verrou KM, Stellos K, Sfikakis PP, Paraskevis D. The role of A-to-I RNA editing in infections by RNA viruses: Possible implications for SARS-CoV-2 infection. Clin Immunol. 2021 May;226:108699. doi: 10.1016/j.clim.2021.108699. Epub 2021 Feb 25. PMID: 33639276; PMCID: PMC7904470.

28. Cherian S, Potdar V, Jadhav S, Yadav P, Gupta N, Das M, Rakshit P, Singh S, Abraham P, Panda S, Team N. SARS-CoV-2 Spike Mutations, L452R, T478K, E484Q and P681R, in the Second Wave of COVID-19 in Maharashtra, India. Microorganisms. 2021 Jul 20;9(7):1542. doi: 10.3390/microorganisms9071542. PMID: 34361977; PMCID: PMC8307577.

29. Kannan SR, Spratt AN, Cohen AR, Naqvi SH, Chand HS, Quinn TP, Lorson CL, Byrareddy SN, Singh K. Evolutionary analysis of the Delta and Delta Plus variants of the SARS-CoV-2 viruses. J Autoimmun. 2021 Aug 11;124:102715. doi: 10.1016/j.jaut.2021.102715. Epub ahead of print. PMID: 34399188; PMCID: PMC8354793.

30. McCallum M, Walls AC, Sprouse KR, Bowen JE, Rosen L, Dang HV, deMarco A, Franko N, Tilles SW, Logue J, Miranda MC, Ahlrichs M, Carter L, Snell G, Pizzuto MS, Chu HY, Van Voorhis WC, Corti D, Veesler D. Molecular basis of immune evasion by the delta and kappa SARS-CoV-2 variants. bioRxiv [Preprint]. 2021 Aug 12:2021.08.11.455956. doi: 10.1101/2021.08.11.455956. PMID: 34401880; PMCID: PMC8366796.

31. Jaafar R, Boschi C, Aherfi S, Bancod A, Bideau ML, Edouard S, Colson P, Chahinian H, Raoult D, Yahi N, Fantini J, La Scola B. High individual heterogeneity of neutralizing activities against the original strain and 9 different variants of SARS-CoV-2. 2021 doi: 10.21203/rs.3.rs-783298/v1

32. Planas D, Veyer D, Baidaliuk A, Staropoli I, Guivel-Benhassine F, Rajah MM, Planchais C, Porrot F, Robillard N, Puech J, Prot M, Gallais F, Gantner P, Velay A, Le Guen J, Kassis-Chikhani N, Edriss D, Belec L, Seve A, Courtellemont L, Péré H, Hocqueloux L, Fafi-Kremer S, Prazuck T, Mouquet H, Bruel T, Simon-Lorière E, Rey FA, Schwartz O. Reduced sensitivity of SARS-CoV-2 variant Delta to antibody neutralization. Nature. 2021 Aug;596(7871):276–280. doi: 10.1038/s41586-021-03777-9. Epub 2021 Jul 8. PMID: 34237773.

33. Laiton-Donato K, Franco-Muñoz C, Álvarez-Díaz DA, Ruiz-Moreno HA, Usme-Ciro JA, Prada DA, Reales-González J, Corchuelo S, Herrera-Sepúlveda MT, Naizaque J, Santamaría G, Rivera J, Rojas P, Ortiz JH, Cardona A, Malo D, Prieto-Alvarado F, Gómez FR, Wiesner M, Martínez MLO, Mercado-Reyes M. Characterization of the emerging B.1.621 variant of interest of SARS-CoV-2. Infect Genet Evol. 2021 Aug 14;95:105038. doi: 10.1016/j.meegid.2021.105038. Epub ahead of print. PMID: 34403832; PMCID: PMC8364171.

